# Evaluating the N1-P2 interpeak latency of the eCAP and its inter-trial variability as potential indicators of neural synchrony in the cochlear nerve of cochlear implant users

**DOI:** 10.1101/2025.05.01.25326811

**Authors:** Shuman He, Ian C. Bruce, Zi Gao, Ross A. Aiello, Christopher R. Mueller

## Abstract

**Objective:** This study evaluated interpeak latency (IPL) and its inter-trial variability (VIL) of the electrically evoked compound action potential (eCAP) as potential alternatives to the phase-locking value (PLV) for quantifying cochlear nerve (CN) synchrony in cochlear implant (CI) users.

**Design:** The IPL was assessed in postlingually deafened adults and three pediatric populations: children with auditory neuropathy spectrum disorder, cochlear nerve deficiency, and typical sensorineural hearing loss. VIL was evaluated only in adults. Their associations with temporal resolution and speech perception outcomes were evaluated. Frequency analysis was conducted to understand the impacts of eCAP recording noise on IPL, VIL, and PLV. Simulations of inter-trial jitter in the eCAP were performed to quantify how the IPL, VIL, and PLV metrics varied with increased temporal jitter.

**Results:** eCAP traces recorded in all patient groups showed a multi-peak issue affecting the accuracy of IPL and VIL assessments. Temporal resolution and speech perception outcomes were significantly correlated with VIL but not with IPL metrics. The PLV was impacted less by recording noise than either the IPL or the VIL. Simulation results revealed that the IPL was less sensitive to the amount of inter-trial jitter in the eCAP than were the VIL and the PLV.

**Conclusions:** The IPL is not a reliable indicator of CN synchrony. The VIL is indicative of neural synchrony in the CN but is affected more by the eCAP recording noise than the PLV. The PLV is therefore the preferred measure for quantifying neural synchrony in the CN in CI users.

**Statements and Declarations:** *Conflict of Interest:* None.

*IRB information:* The data reported in this study were collected for the projects that were approved by the biomedical Institutional Review Board (IRB) of The Ohio State University (IRB study #: 2017H0131, 2018H0344 and 2018N0005; PI: Shuman He), and the IRB of the University of North Carolina at Chapel Hill (IRB study #: 12–1737; PI: Shuman He).

*Author Contributions:* SH designed this study, participated in data analysis, drafted and approved the final version of this paper. ICB participated in study design and data analysis, conducted computational modeling work, drafted and approved the final version of this paper. ZG participated in data analysis, provided critical comments, and approved the final version of this paper. RAA participated in data analysis and approved the final version of this paper. CRM participated in data collection and approved the final version of this paper.

*Data Availability Statement:* The data that support the findings of this study are available from the authors upon reasonable request with permissions from The Ohio State University and the University of North Carolina at Chapel Hill.

## INTRODUCTION

The ability of human listeners to detect, discriminate and recognize complex auditory signals depends on temporally organized neural activity in the auditory system that is time-locked to the auditory input (i.e., neural synchrony) [e.g., [1–9]]. For cochlear implant (CI) users, the cochlear nerve (CN) is the first neural structure to receive electrical stimulation from the device. Consequently, the fidelity with which the CN encodes and processes this stimulation critically influences both the quality and quantity of auditory information conveyed to higher-order auditory centers. In animal models, the discharge patterns of CN fibers have been shown to play a key role in encoding temporal cues [10–12]. Consistent with these findings from single-fiber recordings, simulation studies using computational models have demonstrated that poor neural synchrony in the CN results in a degraded neural representation of temporal envelope cues, leading to deficits in the perception of these cues [1,2]. Given the limited availability of temporal fine structure cues and the reduced spectral resolution provided by current CIs [13,14], CI users primarily depend on temporal envelope cues for speech understanding [e.g., [15,16]]. Consequently, neural synchrony in the CN is expected to play a critical role in determining speech perception outcomes in this population. However, the lack of non-invasive methods for assessing CN neural synchrony has historically hindered its investigation in CI users.

Recently, He et al. [17] reported the use of the phase-locking value (PLV) to assess neural synchrony in the CN in CI patients. In this method, the PLV is calculated based on trial-to-trial phase coherence in the summated activity of CN fibers evoked by electrical stimulation (i.e., the electrically evoked compound action potential [eCAP]). Their findings demonstrated the feasibility of applying this method in both adult and pediatric CI populations [17–19], including children with auditory neuropathy spectrum disorder (ANSD). Moreover, consistent with the literature on acoustic hearing [1,2], PLV was found to correlate with temporal resolution (He et al. 2024b) and speech perception in noise [17,18] in postlingually deafened adult CI users. These results suggest that PLV holds strong promise as an objective indicator of perceptual outcomes in CI users. However, as noted by He et al. [17], this method is affected by the sampling rate of CI devices, which varies substantially across CI manufacturers. The extent to which this variability affects PLV measurements remains unclear, posing a challenge for studies involving participants with different CI systems.

As part of the effort to develop a noninvasive method for assessing neural synchrony in the CN in human CI users, we evaluated, in addition to the PLV, the inter-trial variability of the eCAP interpeak latency (VIL) as a potential alternative indicator of neural synchrony [20]. More recently, Schvartz-Leyzac et al. [21] proposed that the interpeak latency (IPL) of the average eCAP waveform could potentially serve as a marker of neural synchrony. Both indicators are based on the morphological characteristics of the eCAP. As a result, their computation does not depend on manufacturer-specific parameters. Therefore, if the validity of the IPL and the VIL as indicators of CN neural synchrony is established, they can effectively address a key limitation of the PLV method.

In the present study, the IPL was defined as the difference between the N1 and P2 latencies of the eCAP waveform for a given electrode in each participant. The inter-trial variability of the IPL (VIL) was defined as the standard deviation of the IPL across 400 eCAP sweeps (i.e., trials) measured at the same electrode in the same participant. The selection of these two metrics was guided by findings in the acoustic hearing literature, which demonstrate that decreased neural synchrony is associated with delayed response peaks and increased inter-trial variability [22–26]. For healthy spiral ganglion neurons (SGNs), spikes elicited by direct electrical stimulation at the single-unit level exhibit considerably less temporal jitter compared to those evoked by synaptic inputs from hair cells (Miller et al. 1999; Miller et al. 2003; Miller et al. 2001). However, axonal dystrophy and demyelination of SGNs – a common issue observed in CI users [e.g., [27–33]] – alter many neural properties [e.g., [34,35]]. These alternations lead to complex changes in spike timing (Resnick et al. 2018), action potential initiation sites [e.g., [36–39]], and slowed action potential conduction. As a result, spike synchronizations within individual CN fibers across repeated stimulations as well as among different CN fibers are reduced, potentially resulting in delayed response peaks and increased inter-trial variability of neural responses during electrical hearing. This scientific basis supports the use of the VIL to quantify temporal jitter of individual CN fibers, as measured using single-fiber recordings in animal models, where smaller VIL values indicate less temporal jitter and thus better neural synchrony [40]. However, as shown by simulation results reported in Resnick et al.[39], the effects of demyelination on fiber sensitivity and spike timing at the single-unit level are complex and highly heterogeneous. Furthermore, these effects strongly depend on the degree of demyelination, which has been shown to vary substantially along the cochlea, even within individual CI patients [33]. As a near-field recorded compound response in CI patients, the eCAP is not only affected by neural factors (e.g., neural synchrony, discharge rate of individual CN fibers, neural survival of the CN) but also affected by non-neural factors (e.g., impedance, electrode-to-modiolus distance). This distinction is important compared to the far-field recorded compound action potential in listeners with acoustic hearing. Therefore, rather than directly extrapolating findings from acoustic hearing to CI patients, it is essential to empirically evaluate whether reduced spike synchronization observed at the single-unit level translates into measurable changes in IPL and VIL in human CI users. This evaluation is a critical, scientifically rigorous prerequisite before applying these metrics in clinical and research contexts involving CI populations.

Evaluations were conducted by 1) comparing results across four patient populations with different pathophysiological conditions affecting the CN, and 2) assessing the associations between the IPL, the VIL, temporal resolution, and speech perception outcomes in CI users. The rationale for this approach is that a robust indicator needs to be sensitive to the pathophysiological condition it probes and predicts the resulting auditory perception outcomes. The first patient population consisted of children with ANSD, a condition characterized by poor neural synchrony that cannot be fully compensated for by the enhanced neural synchronization provided by electrical stimulation [1,41]. The second patient population included children with cochlear nerve deficiency (CND), a group with substantially fewer but otherwise healthy SGNs compared to children with idiopathic sensorineural hearing loss (SNHL) and normal-sized CNs [42,43]. The third patient population was children with typical SNHL. The fourth patient population was postlingually deafened adult CI users, who exhibit greater loss of SGN somas and peripheral processes at the cochlear base relative to the apex [32,44].

Data from all four patient populations were analyzed to evaluate the IPL. Given the robust correlation between the VIL and the PLV revealed by our pilot results [20], VIL analyses were restricted to the adult CI user dataset. Additionally, relationships between IPL and age, as well as between IPL, VIL, and PLV, were examined in the adult cohort. A frequency analysis of eCAP recording noise was performed to determine how the noise spectrum impacts the frequency-domain PLV metric compared to the time-domain IP and VIL measures. Finally, computational modeling was employed to understand the effect of the amount of temporal jitter on IPL, VIL, and PLV metrics. Based on the literature reviewed previously, shorter IPLs, smaller VILs, and larger PLVs were expected to be indicative of stronger neural synchrony in the CN.

We hypothesized that reduced spike synchronization within and across CN fibers would lead to greater inter-trial variability of the eCAP, which can be quantified using the VIL. This study also tested the hypothesis proposed by Schvartz-Leyzac et al. [21] regarding the effect of reduced neural synchrony on the IPL. Based on these hypotheses, children with typical SNHL were expected to show shorter IPLs than children with ANSD and adult CI users. Among adult CI users, longer IPLs and larger VILs were predicted at basal electrode locations compared to more apical sites. Furthermore, longer IPLs and larger VILs were anticipated to correlate with poorer temporal resolution and lower speech perception scores in noise. Finally, strong correlations were expected among IPL, VIL, and PLV metrics measured within the same patient cohort.

## MATERIALS AND METHODS

### Datasets

These evaluations utilized three existing datasets collected across five studies [17,18,41,45,46], along with one new dataset obtained from a subgroup of 14 adult CI users (A003L, A008R, A009L, A020R, A024R, A035L, A052L, A061R, A070R, A075R, A077L, A080L, A091L, and A093R) who also participated in the study reported in Gao et al. [18]. For the first dataset, the eCAP was recorded as the averaged response across 50 sweeps using a system gain of 50 dB, which are the default parameters used in Custom Sound EP software (Cochlear Ltd, Macquarie, NSW, Australia). In the second dataset, the eCAP was recorded as the averaged response across 100 sweeps with a system gain of 40 dB. It has been shown that the number of averaged sweeps needs to be doubled (i.e., 100 sweeps) when the system gain is set at 40 dB to preserve the required signal quality for eCAP recordings [47]. For the third and fourth datasets, the recorded eCAP response was 400 single sweeps with a default system gain of 50 dB.

The first dataset included eCAPs evoked by single-pulse stimulation measured at three electrode locations across the electrode array (default electrodes, listed from basal to apical locations: electrodes 3, 12, and 19) measured in each of the 30 ears tested in 24 children with ANSD and in each of the 29 ears in 26 children with typical SNHL. These patients participated in the research study comparing temporal response properties of the CN that has been reported in He et al. [45]. The results measured in the right ear of one child with SNHL (S1) were not included due to data file corruption. A subgroup of 10 children with ANSD also participated in a research study assessing the association between objective within-channel gap detection threshold (GDT) measured at electrode 12 and the Phonetically Balanced Kindergarten (PBK) word score (He et al. 2013).

The second dataset included eCAPs measured at the maximum comfortable level using biphasic pulses with an interphase gap (IPG) of 7 µs at one basal, one middle and one apical electrode locations in 30 children with CND. For each participant, only one ear was tested. These children participated in a study evaluating the effect of increasing the IPG of biphasic pulses on eCAPs [46].

The third dataset included eCAP results and AzBio sentence scores measured in quiet and in a 10-talker babble noise with signal-to-noise ratios (SNRs) of +10 and +5 dB in 28 postlingually deafened adult CI users. For each participant, 400 eCAP sweeps were recorded at the maximum comfortable level using biphasic pulses with an IPG of 7 μs at four electrode locations across the electrode array (default electrodes, listed from basal to apical locations: electrodes 3, 9, 15, and 21). The IPL was calculated based on the average responses of the 400 sweeps, and the VIL was calculated based on the latencies of individual sweeps, as detailed in the next section. Psychophysical within-channel GDTs were measured at two electrode locations in each of a subgroup of 15 adult participants. These adult patients participated in two of our previous studies [17,18].

The fourth (i.e., the new) dataset included eCAPs measured at 0 clinical units with an extended recording window to include 64 samples (∼3,200 μs) in a subgroup of 14 adult CI users. The default recording window implemented in Custom Sound EP for measuring the eCAP includes 32 samples (∼ 1600 μs). All other eCAP stimulating and recording parameters, as well as the electrode locations tested were the same as those used in the third dataset for the same group of patients. These extended-window data were utilized exclusively in a frequency analysis to assess the likely impact of eCAP recording noise on IPL, VIL and PLV.

All study participants used a Cochlear™ Nucleus® device (Cochlear Ltd, Macquarie, NSW, Australia) in the test ear. Demographic information for all participants, the electrode locations tested in these experiments, as well as detailed procedures for measuring the eCAP, the PLV, GDT, and speech perception outcomes have been reported in our studies cited above. Therefore, these pieces of information were not included here to avoid redundancy.

### Calculation of the IPL and the VIL

As illustrated in panels (a) (d) of Figure 1 and panel (a) of Figure 6, both single-sweeps and averaged eCAP waveforms frequently exhibited multiple sampled points within the designated latency windows for the N1 and P2 peaks that had nearly identical minimum or maximum voltage values. The time windows containing these points are highlighted in the bolded black rectangles. Closer inspection revealed that the voltage differences among these points were less than 0.5 μV, substantially smaller than the estimated recording noise for Cochlear™ Nucleus® devices, which is approximately 2–5 μV [48–50]. However, the latency difference between these sampled points was 97.6 μs (i.e., two samples) or greater. Consequently, this “multi-peak” issue introduced variability in the calculation of IPL and its VIL, depending on which sampled point was selected to represent the peak latency.

**Figure 1.**
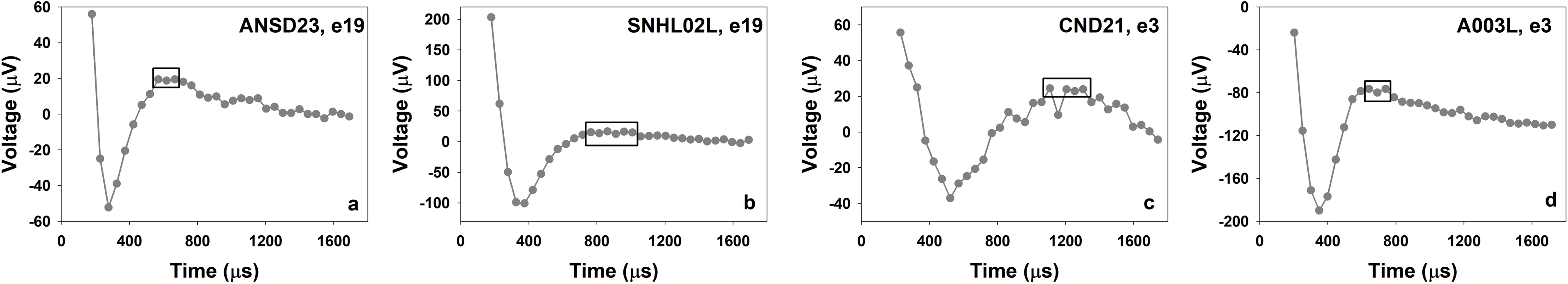
Examples of averaged eCAP response recorded in one patient from each patient group with the multi-peak issue. The time windows where multiple sampled points showed the same values are indicated using rectangles.

Traditionally, N1 and P2 latencies have been determined by identifying the minimum and maximum voltage values within designated latency windows or sample points [51]. Using this approach, single N1 and P2 peaks are identified even when the voltage differences between neighboring samples are as small as 0.0001 μV (i.e., the smallest measurable voltage difference using the setup described in the current study), despite the fact that these small voltage differences are well below the noise floor of CI devices and therefore not clinically meaningful. As a result, the presence of “multi-peak” issues in eCAP waveforms and their impact on latency estimation have not been well documented or systematically addressed. To mitigate this issue, we applied a peak-picking method that took this issue into consideration to a certain extent. Specifically, N1 and P2 latencies were identified within defined latency windows (150–600 μs for N1; 400–1000 μs for P2) by first rounding all voltage values to the nearest integer (in μV) prior to peak selection. This rounding introduced an average voltage resolution of ∼0.5 μV and ensured that the extreme values within a peak latency window differing by less than 1 μV would be considered equivalent. This threshold is more conservative than the lowest noise floor reported for Cochlear™ Nucleus® eCAP recordings (∼ 2 μV) [48] and therefore served as a conservative estimation for the impact of the multi-peak issue on study results. The IPL was defined as the difference between these two peak latencies (i.e., P2 minus N1). For traces with the multi-peak issue, the shortest IPL (IPL_short_) and longest IPL (IPL_long_) were measured. The IPL_short_ was calculated as the difference between the shortest P2 latency and the longest N1 latency, and the IPL_long_ was calculated as the difference between the longest P2 latency and the shortest N1 latency. For traces without the multi-peak issue, the same IPL value was used as the IPL_short_ and IPL_long_ in the subsequent analysis. The variabilities of the IPL_short_ (VIL_short_), the IPL_long_ (VIL_long_), as well as the IPL averaged across all possible N1-P2 pairs (VIL_all_), were defined as the standard deviation of the corresponding IPL values of the 400 eCAP single sweeps recorded at each electrode location in each adult participant.

To compare IPLs across patient groups, IPLs were calculated for eCAPs recorded in three pediatric patient groups and the averaged eCAP across the first 50 sweeps measured in adult CI users. The IPL of the averaged eCAP response across all 400 single-sweep eCAPs was used to evaluate associations with psychophysical GDT, AzBio sentence scores measured in different listening conditions, the PLV, and age in adult CI users. Separate analyses were performed for both the IPL_short_ and IPL_long_, as defined in the previous section. To calculate the VIL, the IPL for each of the 400 single-sweep eCAPs recorded in adult CI users was calculated, allowing for evaluation of variability across trials.

### Statistical Analyses

All statistical analyses were conducted separately for the IPL_short_ and IPL_long_. Linear mixed-effects models (LMMs) with patient group and electrode location as fixed effects were used to compare IPLs at three electrode locations between children with ANSD and children with typical SNHL, as well as to compare IPLs at one basal and one apical electrode location across all four patient groups included in this study. LMMs with electrode location as fixed effect were used to compare IPLs and VIL_all_ values across electrode locations for adult CI users. All LMMs used a correlated regression model with an unstructured correlation matrix to account for repeated observations per participant. The two ears of bilateral users, if both tested, were treated as separate participants. Estimations were obtained using restricted maximum likelihood with Satterthwaite degrees of freedom. Bonferroni correction was used to adjust for multiple comparisons.

Spearman rank correlation tests were used to assess the associations between the IPL and objective GDT and between the IPL and PBK word scores measured in 10 children with ANSD, as well as between PLVs and IPLs in adult CI users. A Pearson correlation test was used to assess the associations between the VIL_all_ and the PLV. To better compare our results with those reported in Schvartz-Leyzac et al. [5], a Pearson correlation test was also used to evaluate the association between participant age and the IPL at different electrode locations in adult participants. For these tests, Bonferroni correction was used to adjust for multiple comparisons as needed.

Multiple linear regressions (MLRs) with AzBio score as the outcome and either the IPL or the VIL_all_ as the predictor were performed to evaluate the association between speech perception outcomes and each of these two parameters under each testing condition (quiet, +10 dB, and +5 dB SNR). The regression model included participant’s age as a covariate to control its potential effects on speech perception and/or eCAP measurements.

The MLRs were performed in R v4.4.1 (R Core Team 2024). All other statistical analyses for this study were performed using SPSS Statistics (v. 28.0.0) (IBM, Armonk, NY). Statistical significance was determined at the 95% confidence level (i.e., *p* <. 05).

### Recording Noise Analysis

For each electrode tested in each participant in the fourth dataset (i.e., the 0 clinical unit recordings), the single-sweep eCAP recording noise frequency spectrum averaged across 400 trials was calculated using the fast Fourier transform. The percentages of recording noise power within and below the frequency range of the PLV analysis (0.789 to 4.73 kHz) were computed.

### Simulation of the Effects of Inter-trial eCAP Jitter on the IPL, VIL, and PLV

Figure 2 of Schvartz-Leyzac et al. [21] illustrates how inter-trial jitter in the single-sweep eCAP waveform can lead to smearing out of the average eCAP waveform and a lengthening of the IPL of the average waveform. However, they only considered two extreme cases in their schematic illustration to justify the use of the IPL as a measure of neural synchrony. Specifically, the case shown in their Figure 2A has a very small amount of jitter such that the average waveform is very similar to the template single-sweep waveform. In comparison, the second case, shown in their Figure 2B has a much larger amount of jitter such that the N1 peaks for some sweeps overlap with the P2 peaks on other sweeps, which leads to the smearing of the average eCAP waveform. It is unclear how much of this smearing of the average eCAP waveform occurs in cases with intermediate amounts of jitter and thus how sensitive the IPL of the average waveform is to various degrees of neural synchrony. To systematically investigate this issue, we have extended and modified the illustration of Schvartz-Leyzac et al. [21] in the following ways.

First, Schvartz-Leyzac and colleagues implemented a horizontal shift in their jitter simulation, but this leads to an identical single-sweep IPL for each trial. In our simulations we have implemented the jitter as a horizontal linear scaling of the template eCAP waveform, such that the single-sweep IPL varies directly as a function of the scaling factor. This scaling-based approach to applying temporal jitter also leads to the P2 latency having a larger standard deviation than then N1 latency, consistent with the results of our latency analysis for the third dataset.

Second, we analyzed the IPL distributions for this dataset and found that they were generally well fit by a gamma distribution. Panel (a) of Figure 2 shows the histogram of single-sweep IPLs for an example electrode, with the fitted gamma distribution probability density function (pdf) shown by the red curve. We used the function `fitdist` from the Matlab Statistics and Machine Learning Toolbox to fit a gamma distribution pdf to the single-sweep IPL data for each electrode from each participant in the third dataset. Outliers were excluded using the Matlab function `isoutlier`. Panels (b) and (c) of Figure 2 show the resulting histograms of the gamma distribution parameters *a* and *b*, respectively. The median values of the gamma distribution parameters were found to be *a* = 18.6 and *b* = 20.4.

Third, we designated the gamma distribution pdf with these median parameter values to be our baseline single-sweep IPL distribution for our simulations. Alternative gamma distributions with different amounts of jitter were then created by scaling the baseline pdf around its mean by a *jitter factor*, such that a jitter factor of 0.5 produces a single-sweep IPL standard deviation half that of the baseline pdf, while a jitter factor of 2 produces a standard deviation twice that of the baseline distribution. Jitter factors were tested from 0 to 2 in steps of 0.1, and ten simulations were run at each value of the jitter factor. Panel (d) of Figure 2 shows pdfs of a subset of the gamma distributions that were used in the inter-trial jitter simulations. For each run, 400 stimulus repetitions (trials) were implemented by generating a random number drawn from the appropriate gamma distribution using the function `gamrnd` from the Matlab Statistics and Machine Learning Toolbox, and applying the jittered linear scaling of the template eCAP waveform via the Matlab function `interp1`.

Fourth, we created a template eCAP waveform for the simulations based on the average eCAP waveform from electrode 21 of participant A008R, as this electrode had the highest PLV value from the third dataset and a high SNR — see Fig. 2 of He et al [17]. A smoothed version of this waveform was generated by upsampling from 20.492 to 500 kHz using piecewise cubic spline interpolation via the Matlab function `interp1`. A linear horizontal scaling was then applied via `interp1` so that the waveform’s IPL matched the mean IPL from the gamma distribution pdfs shown in panel (d) of Figure 2. The resulting template eCAP waveform is plotted in panel (e) of Figure 2. In the simulations, jittered single-sweep waveforms were excluded as outliers if the N1 peak did not occur within the analysis window, consistent with the methodology used for the data analysis.

**Figure 2.**
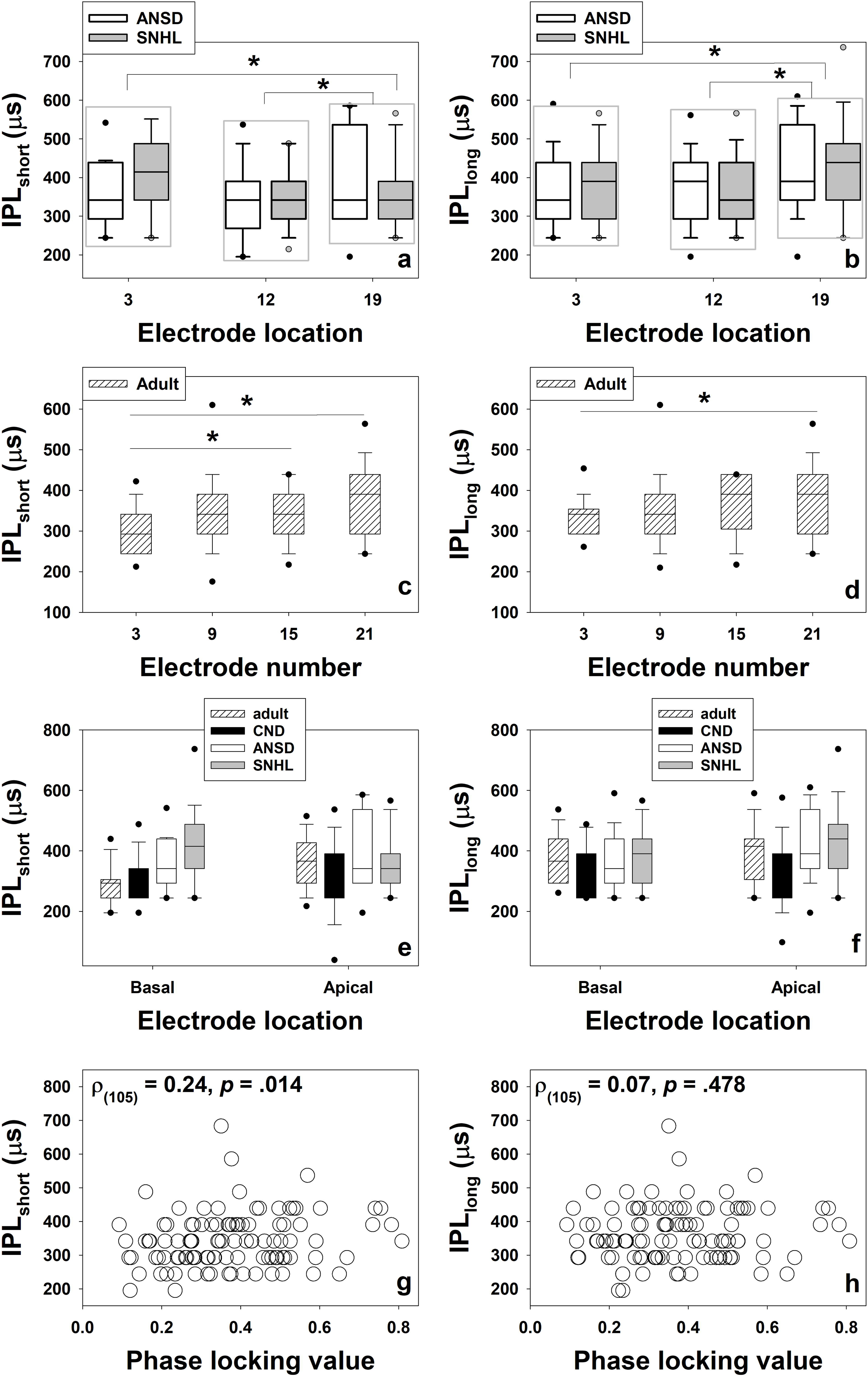
Methodology for simulation of eCAP inter-trial jitter. Panel (a) shows the histogram of single-sweep IPLs for an example electrode. This distribution is well fit by a gamma distribution with the probability density function (pdf) shown by the red curve. Panels (b) and (c) show histograms of the gamma distribution parameters *a* and *b*, respectively, from fits to each electrode in each adult participant from dataset three. Panel (d) shows pdfs of example gamma distributions that are used in the inter-trial jitter simulations. The thick black curve shows the baseline distribution (jitter factor = 1.0) obtained from the median values of *a* and *b* from the histograms in panels (b) and (c). The other gamma distribution pdfs are obtained by scaling the baseline pdf around its mean by the value of *jitter factor*. Panel (e) shows the template eCAP waveform that is used for the inter-trial jitter simulations.

## RESULTS

### Evaluations of the IPL

Using a voltage rounding threshold of 1 μV, the multi-peak issue was observed in 22 out of 86 (25.58%), 15 out of 82 (18.29%), 15 out of 93 (16.13%), and 39 out of 107 (36.44%) eCAPs recorded in children with ANSD, children with SNHL, children with CND and adult CI users, respectively. The difference between the IPL_short_ and IPL_long_ ranged between 48.80 μs (one sample) and 146.40 μs (three samples) (mean: 86.50 μs, SD: 33.44 μs), between 48.80 μs (one sample) and 341.60 μs (seven samples) (mean: 91.50 μs, SD: 79.44 μs), between 48.8 μs (one sample) and 195.2 μs (four samples) (mean: 103.70 μs, SD: 58.76 μs), and between 48.8 μs (one sample) and 146.4 μs (three samples) (man: 58.56 μs, SD: 25.20 μs) for children with ANSD, children with SNHL, children with CND, and adult CI users, respectively. These findings demonstrate that the multi-peak issue introduces variability in IPL measurements across patient groups and can span a range of up to seven samples (341.60 μs). Detailed summary statistics (range, mean, and standard deviation) the IPL_short_ and IPL_long_ measured at different electrode locations in different patient groups are included in Table 1.

**Table 1.** Ranges (means, *SD*s) of the shortest and the longest interpeak latencies measured at different electrode locations in four patient groups. ANSD, auditory neuropathy spectrum disorder; SNHL, sensorineural hearing loss; CND, cochlear nerve deficiency; IPL, interpeak latency; IPL_short_, the shortest IPL; IPL_long_, the longest IPL.

| Dataset | Patient Group | Electrode Location | IPL<sub>short</sub> (μs) | IPL<sub>long</sub> (μs) |
| --- | --- | --- | --- | --- |
| He et al. (2016) | Children with ANSD | 3 | 244.00 – 585.60 (355.54, 83.85) | 244.40 – 634.40 (372.97, 98.69) |
|  |  | 12 | 195.20 – 585.60 (341.60, 98.47) | 195.20 – 585.60 (370.21, 98.05) |
|  |  | 19 | 195.20 – 585.60 (393.77, 120.20) | 195.20 – 634.40 (413.96, 118.55) |
|  | Children with SNHL | 3 | 244.00 – 585.60 (368.71, 98.84) | 244.00 – 585.60 (377.75, 97.22) |
|  |  | 12 | 195.20 – 488.00 (345.21, 83.35) | 244.00 – 585.60 (374.13, 91.80) |
|  |  | 19 | 244.40 – 780.80 (416.54, 129.78) | 244.00 – 780.80 (432.23, 199.95) |
| He et al. (2020) | Children with CND | Basal | 195.20 – 488.00 (305.39, 73.44) | 244.00 – 488.00 (325.86, 78.07) |
|  |  | Middle | 195.20 – 488.00 (313.25, 74.30) | 195.20 – 536.80 (321.14, 85.25) |
|  |  | Apical | 97.60 – 536.80 (312.32, 106.92) | 195.20 – 634.40 (338.35, 103.27) |
| Gao et al. (2024) | Adult | 3 | 195.20 – 439.20 (304.06, 57.42) | 244.00 – 488.00 (332.22, 51.68) |
|  |  | 9 | 146.40 – 683.20 (339.65, 99.09) | 195.20 – 683.20 (359.17, 94.37) |
|  |  | 15 | 195.20 – 439.20 (343.34, 64.36) | 195.20 – 439.20 (367.74, 73.33) |
|  |  | 21 | 244.00 – 585.60 (371.23, 92.34) | 244.00 – 585.60 (386.91, 93.85) |

### Effects of participant group and electrode location

The results of the IPL_short_ and IPL_long_ measured at three electrode locations in children with ANSD and children with typical SNHL are shown in panels (a) and (b) of Figure 3, respectively. For both sets of results, LMMs revealed no significant effect of patient group (IPL_short_: *F_(1, 54.73)_* = 0.34, *p* = .561; IPL_long_: *F_(1, 54.94)_* = 0.12, *p* = .733), or the interaction between participant group and electrode location (IPL_short_: *F_(2, 55.17)_* = 0.18, *p* = .839; IPL_long_: *F_(2, 54.52)_* = 0.17, *p* = .848) on the IPL. However, there was a significant effect of electrode location on the IPL (IPL_short_: *F_(2, 55.17)_* = 6.88, *p* = .002; IPL_long_: *F_(2, 54.50)_* = 6.06, *p* = .004). Post-hoc pairwise comparisons showed that IPLs measured at electrode 19 were significantly shorter than those measured at both electrode 3 (IPL_short_: *p* = .032; IPL_long_: *p* = .015) and electrode 12 (IPL_short_: *p* = .002; IPL_long_: *p* = .006) across both patient groups. There was no significant difference between IPLs measured at electrodes 3 and 12 (IPL_short_: *p* = .628; IPL_long_: *p* = 1.000).

**Figure 3.**
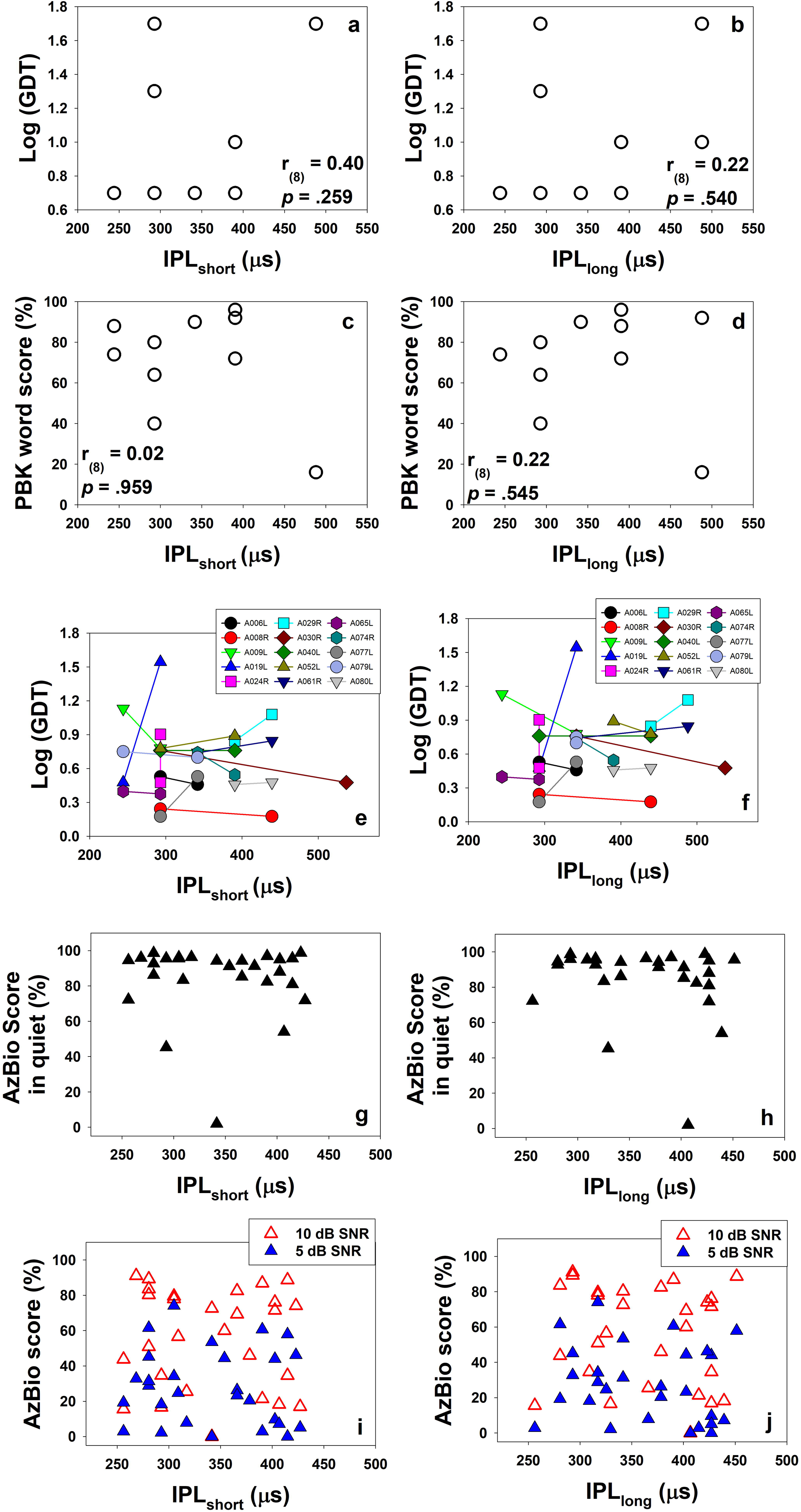
N1-P2 interpeak latencies (IPLs) of the eCAP measured in different patient groups, as well as their associations with phase locking values (PLV). Panels (a) and (b) show box plots of IPL_short_ and IPL_long_ values measured at three electrode locations in children with auditory neuropathy spectrum disorder (ANSD) and children with idiopathic sensorineural hearing loss (SNHL), respectively. Panels (c) and (d) show IPL_short_ and IPL_long_ results measured at four electrode locations in adult participants, respectively. For these four panels, lines and asterisks indicate groups or electrode locations with statistically significant differences. Panels (e) and (f) show box plots of IPL_short_ and IPL_long_ values measured at two electrode locations in all four patient groups, respectively. For all box plots included in this figure, boxes show the range between the first and the third quartiles of the data values. The horizontal bars inside the boxes represent the median. The vertical whiskers show the range of values that are within 1.5 interquartile range (IQR) from the boxes. The dots show the 5th and 95th percentile of the data. Panels (g) and (h) show the association between IPLs quantified in two ways and the PLV measured in adult participants. Each dot indicates the result measured at one electrode location in one participant. The results of Spearman rank correlation tests are indicated in these two panels.

Panels (c) and (d) of Figure 3 show the results of the IPL_short_ and IPL_long_ measured at four electrode locations in adult participants. Overall, the data reveal an increasing trend in IPL as electrode location moves toward more apical sites for both measures. LMMs confirmed this visual observation, showing a significant effect of electrode location on both the IPL_short_ (*F_(3, 25.34)_* = 5.98, *p* = .003) and IPL_long_ (*F_(3, 24.80)_* = 3.65, *p* = .026). Post-hoc paired comparisons indicated that IPL_short_ values measured at electrode 3 were significantly shorter than those measured at electrodes 15 (p = .023) and 21 (*p* = .002). IPL_long_ values measured at electrode 3 were also significantly shorter than those measured at electrode 21 (*p* = .020). No other significant differences between electrode pairs were observed for either measure.

To better understand the significant electrode location effect on the IPL observed in adult participants, we compared IPL_short_ and IPL_long_ values measured at two electrode locations across four patient groups. These results are shown in panels (e) and (f) of Figure 3. For both the IPL_short_ and IPL_long_, LMMs revelated significant effects of patient group (IPL_short_: *F_(3, 111.52)_* = 7.37, *p* < .001; IPL_long_: *F_(3, 111.27)_* = 4.43, *p* = .006) and electrode location (IPL_short_: *F_(1,110.27)_* = 11.99, *p* < .001; IPL_long_: *F_(1,109.57)_*= 9.55, *p* = .003). The interaction of these two factors on the IPL was not statistically significant (IPL_short_: *F_(3, 110.32)_* = 1.29, *p* =.280; IPL_long_: *F_(3, 109.64)_*= 0.80, *p* = .497). Regardless of how the IPL was quantified, IPLs measured at the basal electrode location were significantly shorter than those measured at the apical electrode location for all patient groups (IPL_short_: *p* < 001; IPL_long_: *p* = .002). For the group effects on the IPL_short_ and IPL_long_, post-hoc pairwise comparisons showed slightly different results. Specifically, children with CND had significantly smaller IPL_short_ values than both children with ANSD (*p* = .009) children with typical SNHL (*p* < .001). IPL_short_ values measured in adult CI users were significantly smaller than those measured in children with typical SNHL (*p* = .010). There was no significant difference in the IPL_short_ between any other two patient groups (*p* > .05). For the results of the IPL_long_, children with CND showed significantly shorter IPLs than children with ANSD (*p* = .033) and children with typical SNHL (*p* = .009). There was no significant difference in the IPL_long_ between any other two patient groups (*p* > .05). The lack of significant difference in the IPL_short_ or IPL_long_ between adult CI users and children with CND should be noted and highlighted here due to its importance for data interpretation.

### Correlations with the PLV

Panels (g) and (h) of Figure 3 show the IPL_short_ and IPL_long_ measured in adult participants plotted as a function of the PLV, respectively. The results of Spearman rank correlation tests showed a significant correlation between the PLV and the IPL_short_ (*ρ_(105)_* = 0.24, *p* = .014) with larger PLV values associated with longer IPL_short_ latencies. However, this significant correlation was not observed for the results of the IPL_long_ (*ρ_(105)_* = 0.07, *p* = .478).

### Associations with temporal resolution, speech perception and advanced age

The associations between GDTs and IPLs quantified in both ways measured at electrode 12 in a subgroup of 10 children with ANSD are shown in panels (a) and (b) of Figure 4. Panels (c) and (d) show PBK word scores measured in the same group of patients plotted against the IPL_short_ and the IPL_long_, respectively. These results showed a lack of association between the IPL, regardless of how it was quantified, and either of the two auditory perception outcomes, which is supported by the results of Spearman rank correlation tests listed in each panel.

**Figure 4.**
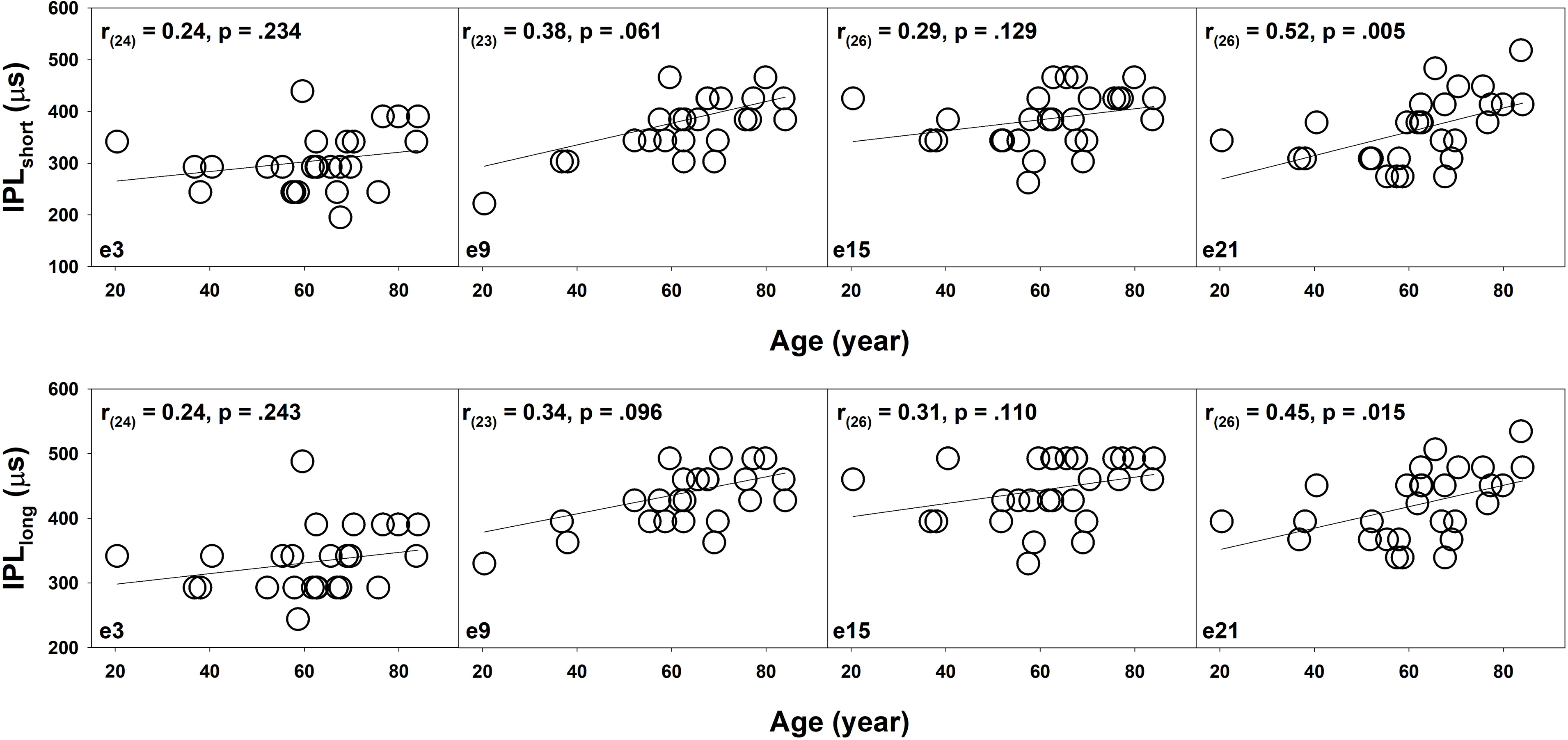
The associations between IPLs quantified in two ways and auditory perception outcomes. Panels listed in the left and right columns represent the results of the IPL_short_ and the IPL_long_, respectively. Panels (a) and (b) show scatter plots of the association between the IPL and objective gap detection threshold (GDT) measured in a subgroup of 10 children with ANSD. Panels (c) and (d) show the Phonetically Balanced Kindergarten (PBK) word score plotted against the IPL for the same group of children with ANSD. The results of Spearman correlation tests are shown in these four panels. Panels (e) and (f) display IPLs and psychophysical GDTs measured at two electrode locations in each of 15 adult participants. Lines connect the data measured at the two electrode locations tested in the same ear. Panels (g) and (h) show scatter plots of the association between the IPL and AzBio sentence scores measured in quiet in adult participants tested in this study. Panels (i) and (l) show scatter plots of the association between the IPL and AzBio sentence scores measured in the two noise conditions in the same group of adult participants. Each symbol indicates the result measured in one participant in these four panels.

Panels (e) and (f) of Figure 4 show psychophysical GDTs measured at two electrode locations per test ear in a subgroup of 15 adult participants as a function of the IPL_short_ and the IPL_long_, respectively. These results clearly demonstrated a lack of consistent trend in the association between the IPL quantified either way and GDT across participants. For the results of the IPL_short_, the two CI electrode locations tested in the right ear of A024 showed equal IPLs but different GDTs. In the left ear of A040, the same GDTs were measured for the two electrode locations with an IPL difference of 97.6 µs (two samples). For the rest of participants, while smaller GDTs were observed at the electrode locations with shorter IPLs in six participants, results measured in the other seven participants showed an opposite pattern. For the results of the IPL_long_, the two CI electrode locations tested in the right ear of A024 and in the left ear of A079 showed equal IPLs but different GDTs. In the left ear of A040, the same GDTs were measured for the two electrode locations with an IPL difference of 146.4 µs (three samples). For the rest of participants, while smaller GDTs were observed at the electrode locations with shorter IPLs in five participants, results measured in the other seven participants showed an opposite pattern. Interestingly, in the left ear of participant A052, the association between GDT and IPL differed depending on how the IPL was quantified—showing opposite patterns for IPL_short_ and IPL_long_. For all other participants, the method used to quantify IPL (i.e., IPL_short_ vs. IPL_long_) did not alter the direction of its relationship with GDT. In contrast, the same set of psychophysical GDT data demonstrated consistent trend of association with PLV values (lower GDTs ∼ higher PLV) across a majority of the participants (He et al. 2024b), with the exception of the data measured in the right ear of A029 and in the left ears of A040 and A080.

Panels (g) (j) of Figure 4 show the associations between the numerically averaged IPL across four electrode locations and AzBio sentence scores measured in quiet and in noise in 28 adult participants, respectively. These data demonstrate a lack of association between the IPL and AzBio sentence scores measured in any listening conditions. This observation was supported by the results of MLRs after controlling for the effect of participant age. Table 2 includes the results of MLRs calculated for IPLs quantified in both ways and AzBio sentence scores measured in different listening conditions.

**Table 2.**
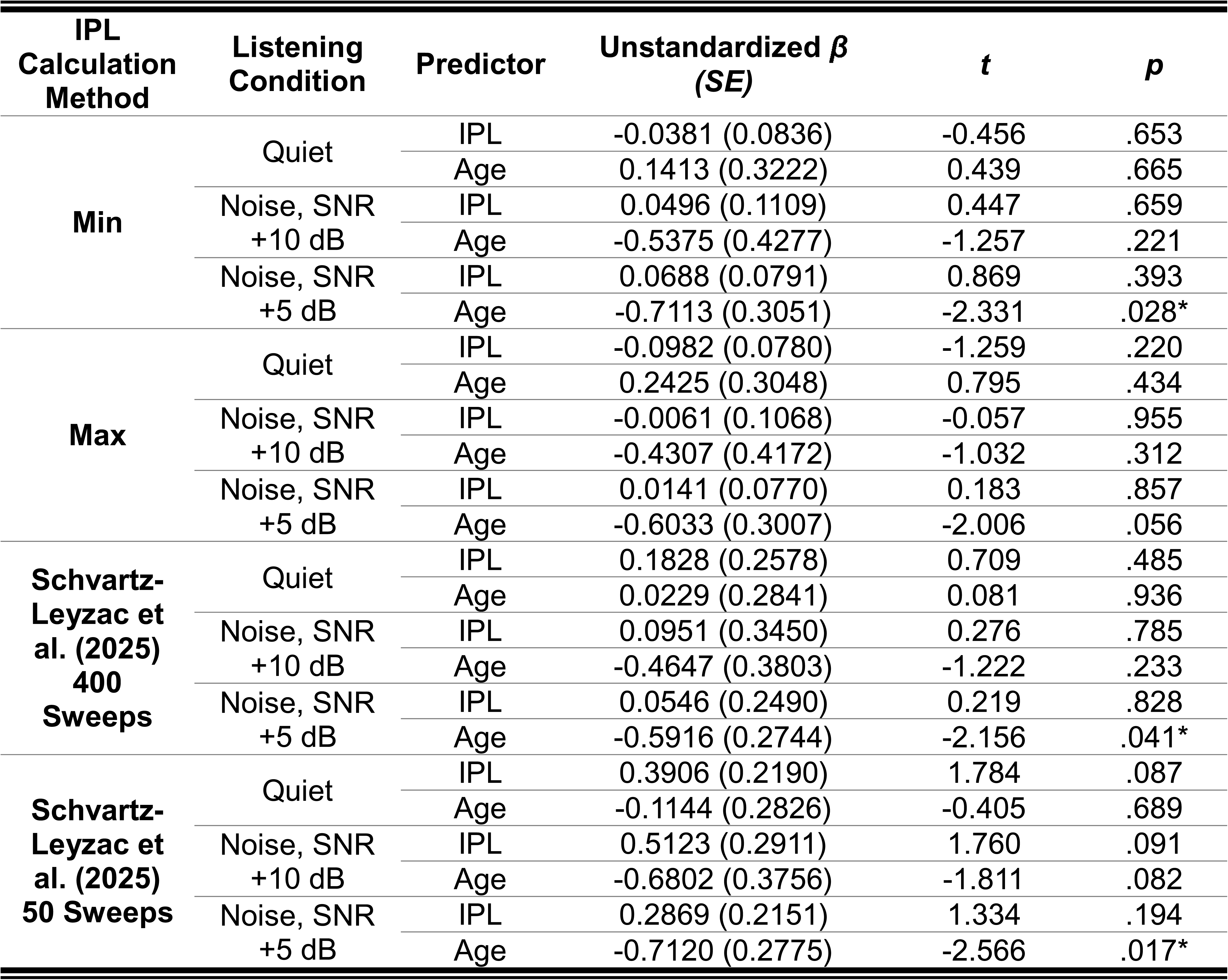
Results of Multiple linear regressions for assessing the associations between AzBio sentence scores measured in different listening conditions and the interpeak latency (IPL) results quantified in different ways after controlling the age effect. SNR: signal to noise ratio; SE: standard error.

| IPL Calculation Method | Listening Condition | Predictor | Unstandardized $\beta$ (SE) | $t$ | $p$ |
| --- | --- | --- | --- | --- | --- |
| **Min** | Quiet | IPL | -0.0381 (0.0836) | -0.456 | .653 |
|  |  | Age | 0.1413 (0.3222) | 0.439 | .665 |
|  | Noise, SNR +10 dB | IPL | 0.0496 (0.1109) | 0.447 | .659 |
|  |  | Age | -0.5375 (0.4277) | -1.257 | .221 |
|  | Noise, SNR +5 dB | IPL | 0.0688 (0.0791) | 0.869 | .393 |
|  |  | Age | -0.7113 (0.3051) | -2.331 | .028* |
| **Max** | Quiet | IPL | -0.0982 (0.0780) | -1.259 | .220 |
|  |  | Age | 0.2425 (0.3048) | 0.795 | .434 |
|  | Noise, SNR +10 dB | IPL | -0.0061 (0.1068) | -0.057 | .955 |
|  |  | Age | -0.4307 (0.4172) | -1.032 | .312 |
|  | Noise, SNR +5 dB | IPL | 0.0141 (0.0770) | 0.183 | .857 |
|  |  | Age | -0.6033 (0.3007) | -2.006 | .056 |
| **Schvartz-Leyzac et al. (2025) 400 Sweeps** | Quiet | IPL | 0.1828 (0.2578) | 0.709 | .485 |
|  |  | Age | 0.0229 (0.2841) | 0.081 | .936 |
|  | Noise, SNR +10 dB | IPL | 0.0951 (0.3450) | 0.276 | .785 |
|  |  | Age | -0.4647 (0.3803) | -1.222 | .233 |
|  | Noise, SNR +5 dB | IPL | 0.0546 (0.2490) | 0.219 | .828 |
|  |  | Age | -0.5916 (0.2744) | -2.156 | .041* |
| **Schvartz-Leyzac et al. (2025) 50 Sweeps** | Quiet | IPL | 0.3906 (0.2190) | 1.784 | .087 |
|  |  | Age | -0.1144 (0.2826) | -0.405 | .689 |
|  | Noise, SNR +10 dB | IPL | 0.5123 (0.2911) | 1.760 | .091 |
|  |  | Age | -0.6802 (0.3756) | -1.811 | .082 |
|  | Noise, SNR +5 dB | IPL | 0.2869 (0.2151) | 1.334 | .194 |
|  |  | Age | -0.7120 (0.2775) | -2.566 | .017* |

Figure 5 shows IPLs measured at different electrode locations in adult participants plotted as a function of age. The upper and lower panels show the results of the IPL_short_ and IPL_long_, respectively. Each panel represents the results measured at one electrode location. Overall, IPLs measured at all four electrode locations showed a trend toward longer latencies with increasing age. However, this trend only reached statistical significance for the IPL_short_ measured at electrode 21, as determined by Pearson correlation tests with Bonferroni correction.

**Figure 5.**
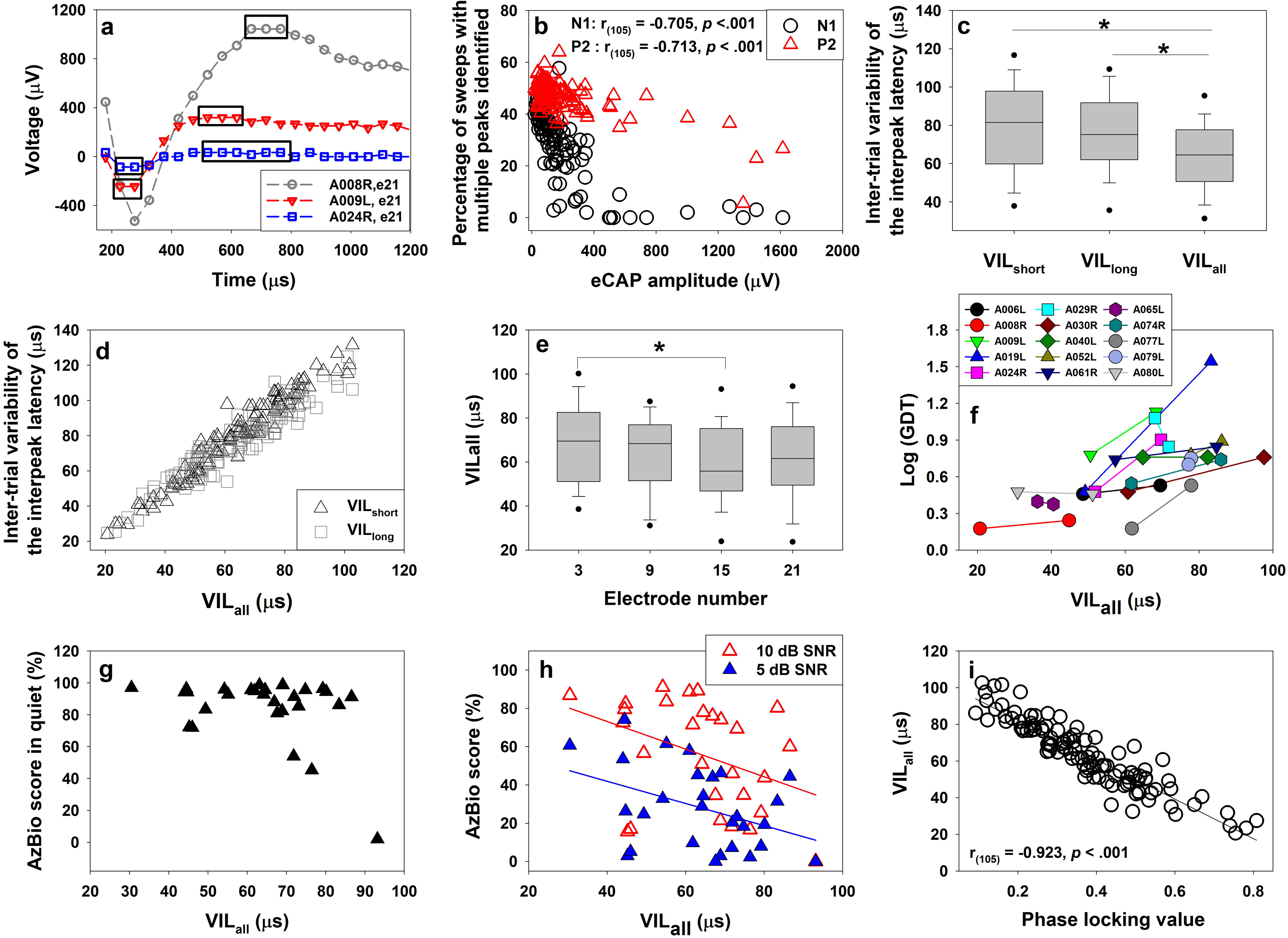
The association between age and the interpeak latency for adult participants. Upper and lower panels show the results of the IPL_short_ and the IPL_long_, respectively. Each panel represents the results measured at one electrode location. Each symbol indicates the result measured in one participant. Solid lines represent the results of linear regression. The results of Pearson correlation tests are indicated in each panel.

### Evaluations of the VIL

Panel (a) of Figure 6 shows example single eCAP sweeps with the multi-peak issue that affects the precision of the VIL. This issue occurred in all participants, even in cases where eCAP amplitude was 1000 µV or larger (i.e., the grey trace with circle marks). The percentage of traces with this multi-peak issue ranges from 0 to 57.65% (mean: 29.13%, SD: 13.93%) and from 5.51 to 63.96% (mean: 46.59%, SD: 7.08%) for the N1 and the P2 peaks, respectively. Panel (b) of Figure 6 shows the percentages of eCAP traces with multiple points identified for the N1 (black circles) and the P2 peaks (red triangles) plotted as a function of eCAP amplitude. These data demonstrate that the percentage decreases with larger eCAP amplitudes for both peaks, which indicates the impact of recording noise.

**Figure 6.**
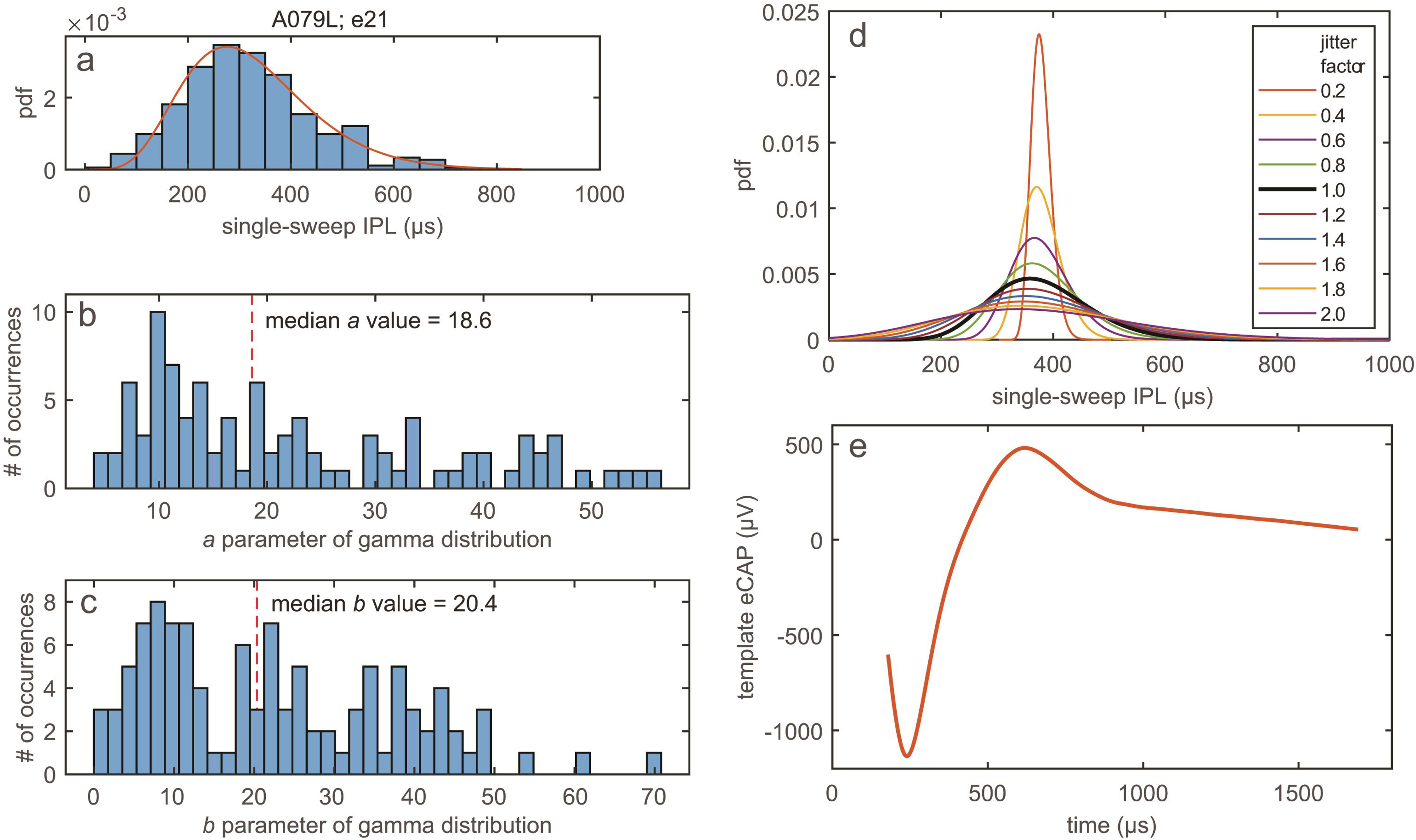
Group results and individual examples of the inter-trial variability of the interpeak latency (VIL) measurement. Panel (a) shows example traces measured at electrode 21 in three participants. The time windows where multiple sampled points showed the same values are indicated using rectangles. Panel (b) shows the percentage of traces showing multiple equal-value points for the N1 (red triangles) and P2 (black dots) plotted against the eCAP amplitude for the results measured at all electrode locations in all participants tested in this study. Panel (c) shows the box plots for VILs calculated based on the longest (VIL_long_) and shortest (VIL_short_) interpeak latencies, as well as for the averaged interpeak latency across all possible interpeak latency values measured for trials with multiple equal-value points (VIL_all_). Panel (d) shows the VIL_long_ and VIL_short_ plotted against VIL_all_. Panel (e) shows VIL_all_ results measured at four electrode locations in adult participants. Panel (f) displays VIL_all_ values and psychophysical GDTs measured at two electrode locations in each of 15 adult participants. Lines connect the data measured at the two electrode locations tested in the same ear. Panels (g) and (h) show scatter plots of the association between the VIL_all_ and AzBio sentence scores measured in quiet and in noise in adult participants tested in this study, respectively. Panel (i) shows the association between the VIL_all_ and the phase locking value measured in adult participants. Boxes displayed in panels (c) and (e) show the range between the first and the third quartiles of the data values. The horizontal bars inside the boxes represent the median. The vertical whiskers show the range of values that are within 1.5 interquartile range (IQR) from the boxes. The dots show the 5^th^ and 95^th^ percentile of the data. Asterisks indicate statistically significant differences. In panels (b), (d) and (h), each symbol represents the result measured at one electrode location in one participant. In panels (g) and (h), each symbol indicates the result measured in one participant. In panels (h) and (i), solid lines represent the results of linear regression. The result of a Pearson correlation test is indicated in panel (i).

Panel (c) of Figure 6 shows the results of the VIL_short_, VIL_long_ and VIL_all_. LMM revealed significant main effect of VIL type, after controlling electrode location (*F_(2, 284.95)_* = 42. 37, *p* < .001). Post-hoc analysis revealed that the VIL_all_ was significantly smaller than both the VIL_short_ and the VIL_long_ (*p* < .001). There was no significant difference between the VIL_short_ and the VIL_long_ (*p* = .117). Due to the strong correlations among these VIL results, as demonstrated by panel (d) Figure 6, the VIL_all_ results were used in the subsequent assessments.

Panel (e) of Figure 6 shows VIL_all_ values measured at four electrode locations in adult participants. These results display an overall decreasing trend in the VIL_all_ as the testing electrode moved to more apical electrode locations. Consistent with this observation, the results of a LMM revealed a significant effect of electrode location on the VIL_all_ (*F_(3, 23.68)_*= 3.56, *p* = .029). Post-hoc analysis revealed that VIL_all_ results measured at electrode 3 were significantly larger than those measured at electrode 15 (*p* = .023). There was no significant difference in the VIL_all_ between any other electrode location pairs (*p* > .05).

The association between psychophysical GDTs and VIL_all_ values is shown in panel (g) of Figure 6. All except for four participants (A029, A040, A065 and A080) showed lower GDTs at the electrode location with smaller VIL_all_ values. The results measured in the right ear of A029 and in the left ears of A065 and A080 showed an opposite pattern. For A040, the same GDTs were measured at the two electrode locations with different VIL_all_ values. Panels (h) and (i) of Figure 6 show the association between the numerically averaged VIL_all_ across four electrode locations and AzBio sentences scores measured in quiet and in noise, respectively. These results demonstrate the lack of association between AzBio sentence scores measured in quiet and the VIL_all_. AzBio sentence scores measured in the two noise conditions were lower for larger VIL_all_ values. These observations are supported by the MLRs after controlling for the effect of age (Quiet: *t_(25)_* = −2.043, *p* = .052; +10 dB SNR: *t_(25)_* = −2.102, *p* = .046; +5 dB SNR: *t_(25)_*= 2.342, *p* = .027).

Finally, panel (i) of Figure 6 shows the VIL_all_ plotted against the PLV. The result of a Pearson correlation test showed a significant correlation between these two parameters (*r_(105)_* = −0.923, *p* < .001), with smaller VILs correlated with larger PLV values.

### Analysis of Recording Noise

Panel (a) of Figure 7 shows the individual (thin lines) and population average (thick black line) recording-noise spectrum for each of 69 electrodes across the 14 adult participants in dataset four. For the population average noise spectrum, only 22.5% of the noise power is within the frequency range of the PLV analysis (0.789 to 4.73 kHz), indicated by the red vertical dashed lines. In contrast, 58% of the population average recording noise power is below this frequency range. Thus, the effective SNR of the PLV measure is substantially better than that of the time-domain IPL and VIL measures which are impacted by the full noise bandwidth. Panels (b) and (c) show histograms for the percentage of the recording noise power that is within the frequency range of the PLV analysis (panel b) and the percentage of power below that frequency range (panel c). Across all the electrodes in dataset four, the percentage of power within the PLV analysis frequency range extends from 10% to 40%, whereas the percentage of power below the PLV analysis range is between 30% and 90%. This indicates that electrodes with a greater amount of total recording noise power than the population average only have slightly more than average noise power within the PLV frequency range but have substantially more noise than the average at lower frequencies. Therefore, the PLV is more robust than the IPL and VIL over a greater range of recording noise powers.

**Figure 7.**
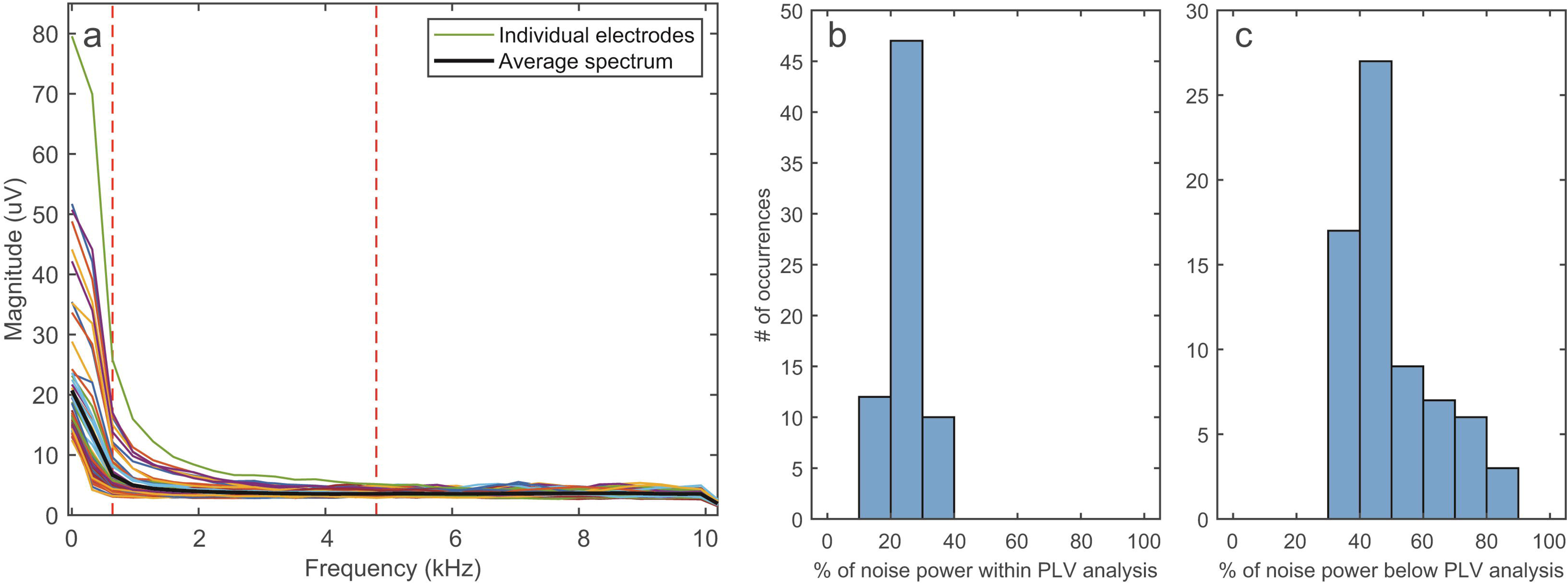
Recording noise analysis. Panel (a) shows the individual (thin lines) and population average (thick black line) recording-noise spectrum for each of 44 electrodes across the 14 adult participants in dataset four. The vertical dashed red lines indicate the frequency range of the PLV analysis (0.789 to 4.73 kHz). Panels (b) and (c) show histograms for the percentage of the total recording noise power that is within the frequency range of the PLV analysis (panel b) and the percentage of power below that frequency range (panel c).

### Simulation of the Effects of Inter-trial eCAP Jitter

The results of the eCAP jitter simulations are shown in Figure 8. Panels (a) (d) provide example results of the eCAP inter-trial jitter simulations, for jitter factors of 0.1, 0.3, 1, and 2, respectively. It can be observed that increasing the jitter factor from 0.1 to 0.3 produces only minimal temporal smearing in the mean eCAP waveform (thick black lines) when compared to the template waveform (thick red dash lines), so the IPL is practically unchanged. When the jitter factor is increased to a value of 1 (i.e., to the baseline gamma distribution pdf that has a median amount of jitter) somewhat more temporal smearing occurs in the mean eCAP waveform and its IPL is elongated compared to the template waveform’s IPL. Increasing the jitter factor to 2 produces substantially more temporal smearing and a greatly lengthened IPL in the mean waveform.

Panels (e) (g) of Figure 8 show the mean values of the estimated IPL of the mean eCAP waveform, the VIL over the 400 trials, and the PLV over 400 trials, respectively, as a function of the jitter factor. Error bars show the standard deviation in the value over ten simulation repetitions. These results clearly show that the IPL is very insensitive to the jitter factor over the range of 0 to around 0.5, and then from 0.5 to 1 the IPL starts increasing and has a roughly linear relationship with the jitter factor for values between 1 and 2. However, the standard deviation of the estimates (indicated by the error bars) also grows in this region, indicating that it lacks somewhat in reliability even within this range of jitter factor values. In contrast, the VIL and the PLV show a nearly linear relationship with the jitter factor over the entire range from 0 to 2, with only a moderate increase in the standard deviation of the estimates at larger jitter factor values. These simulation results strongly suggest that the VIL and PLV are much more reliable measures of CN synchrony than the IPL of the mean waveform.

**Figure 8.**
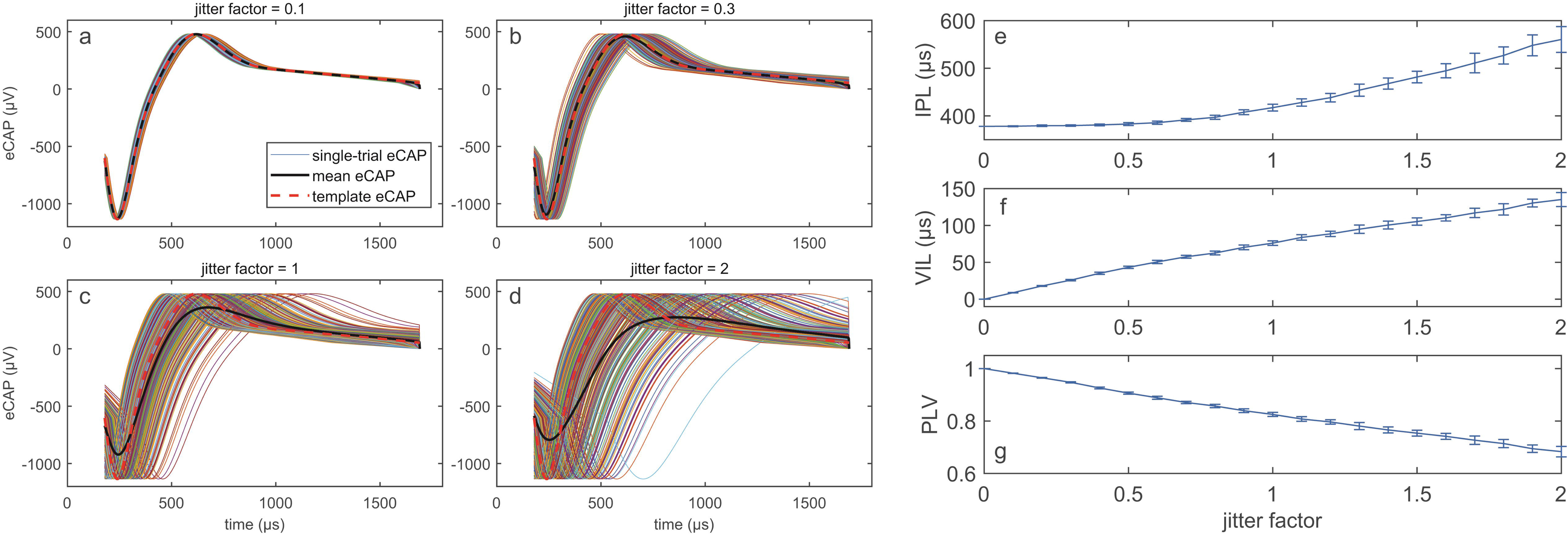
Jitter simulation results. Panels (a) (d) show example results of the eCAP inter-trial jitter simulations, for jitter factors of 0.1, 0.3, 1, and 2, respectively. Thin lines show the eCAP waveforms for single trials, thick black lines show the mean eCAP over all the trials, and thick dashed red lines show the template eCAP waveform. Panels (e) (g) show the estimated IPL of the mean eCAP waveform, the VIL, and the PLV, respectively, as a function of the jitter factor. Each data point is the mean value from ten simulations run at that jitter factor, with error bars showing the standard deviation in the value over the ten repetitions.

## DISCUSSION

This study evaluated the IPL and the VIL as potential indicators of neural synchrony in the CN in CI users. Both metrics require accurately identifying latencies of the N1 and P2 peaks of the eCAP. In this section, two major factors affecting the accuracy of both measures were discussed first, followed by discussions of the results of each assessment.

### Recording noise and sampling rate

In current CIs, the level of recording noise for eCAP measurements varies across manufacturers and is dependent on the number of sweeps averaged to obtain the eCAP. The noise floor is reported to be approximately 20 μV in older for Cochlear™ Nucleus® implants and 2 5 μV in new for Cochlear™ Nucleus® implants [48] and 20 50 μV (Glassman & Hughes 2012) in Advanced Bionics devices. For MED-EL devices, the noise floor is frequency dependent, with an estimated overall noise level of 2.6 10 μV [52,53]. In our analysis of measured recording noise of the fourth dataset, we found that the single-sweep noise amplitudes for these adult Cochlear™ Nucleus® users is also frequency dependent — see panel (a) of Figure 7. As demonstrated by our eCAP results, this recording noise introduces variability in IPL and VIL assessments.

The temporal resolution of peak latency measurements depends on the sampling rate used for eCAP recordings. Higher sampling rates enable more precise identification of response peaks, leading to more accurate IPL and VIL calculations. In Cochlear™ Nucleus® devices, the sampling rate is 20,492 Hz, resulting in a step size (i.e., the time interval between consecutive samples) of 48.8 μs. This relatively large step size can increase both local and global errors in peak latency estimation. Advanced Bionics devices offer a sampling rate of 56 kHz, corresponding to a finer temporal resolution of approximately 17.85 μs. However, the benefit of this higher resolution is offset by device’s relatively high noise floor. Theoretically, the high sampling rate (i.e.,1.2 MHz) and the relatively low noise floors of MED-EL devices could improve the accuracy of IPL and VIL assessments. More studies are warranted to test this theoretical possibility. Overall, these two hardware-related factors—the noise floor and the sampling rate— limit the accuracy of both IPL and VIL metrics.

### Interpeak latency of the eCAP

One aim of this study was to test the hypothesized effect of reduced neural synchrony of the CN on the IPL of the averaged eCAP across 50-100 sweeps, as proposed by Schvartz-Leyzac et al. [21]. Overall, our results do not provide compelling evidence to support the use of IPL as an indicator of neural synchrony in the CN. The key findings supporting this conclusion are summarized and discussed below.

**Finding 1:** There was no significant group difference in the IPL between children with ANSD, a patient population characterized by poor neural synchrony, and children with typical SNHL. This lack of group difference between these two patient populations questions the sensitivity of IPL as a reliable indicator of neural synchrony in the CN in human CI users.
**Finding 2:** In adult CI users, IPLs increased as the testing electrode moved toward more apical locations. If the IPL exclusively reflects neural synchrony in the CN, these results would suggest that adult CI users have better neural synchrony at the base than at more apical regions of the cochlea. However, this interpretation conflicts with histological findings from human temporal bone studies in CI users showing greater neural degeneration at the base than at the apex of the cochlea [32,44], as well as neural degenerated patterns observed in age-related hearing loss [e.g., [54]]. Moreover, children with CND, a patient population with substantially fewer but otherwise healthy SGNs, showed significantly shorter IPLs than both children with ANSD and children with typical SNHL. This suggests that SGN count may affect the IPL, with reduced SGN leading to shorter IPLs. The lack of significant difference in IPLs between adult CI users and children with CND, combined with the well-documented SGN loss in patients with SNHL [27–33,54], further support the potential impact of SGN count on the IPL. Collectively, these findings raise questions about the biological mechanisms that IPL measurements actually reflect.
**Finding 3**. In adult CI users, IPLs measured at all four electrode locations showed an increasing trend with advancing age, with a stronger trend at the apical electrode location. These age-related increases are inconsistent with prior evidence of greater CN degeneration at the cochlear base than apex in older adults [54,55].
**Finding 4**. No significant associations were found between IPL and either temporal resolution or speech perception outcomes in children with ANSD or in adult CI users. These results contradict the established relationship between poor neural synchrony in the CN and deficits in temporal resolution and speech perception in noise [e.g., [1]].
**Finding 5**. Consistent with the experimental data, our simulation results demonstrated IPL assessment’s relative lack of sensitivity to the amount of neural synchrony and vulnerability to high measurement error, which further supported this conclusion.

In summary, our data show that the IPL, as a proposed indicator of neural synchrony in the CN, lacks both sensitivity to the relevant underlying neural pathology and predictive value for the resulting auditory perceptual outcomes. Therefore, the current findings do not support the use of IPL as a valid marker of neural synchrony in the CN in CI users.

#### Study results comparisons

In a recent study, Schvartz-Leyzac et al. [21] proposed that decreased neural synchrony should lead to large IPL values. In their study, a significant moderate correlation between the IPL and participant age in data pooled across all electrodes in all adult participants was found, with longer IPLs correlated with more advanced age (see their Figure 5). Our data, as shown in Figure 5, showed a similar overall pattern, with this positive correlation being stronger at more apical electrode locations. As discussed above, this pattern is not consistent with neural degenerated patterns observed in age-related hearing loss [e.g., [54]]. At this point, factors accounting for this observed association between age and the IPL remain unknown. It should be noted that our simulation results do not exclude the possibility that the IPL of the mean eCAP waveform can be affected by pathologies that cause a deterministic spreading out of the spike times in a population of CN fibers, leading to a common, broadened eCAP waveform across trials. In these cases, longer IPLs will be observed but they are not due to reduced neural synchrony in the CN. We speculate that the IPL is likely affected by both deterministic changes in the eCAP waveform (common across trials) and inter-trial jitter in the eCAP, suggesting that reduced neural synchrony is unlikely to be the only factor underlying the age-related increase in the IPL.

Furthermore, Schvartz-Leyzac et al. [21] reported a negative correlation between AzBio sentence scores at + 10 dB SNR and the IPL (see their Fig. 8), suggesting that longer IPLs were associated with poorer speech perception in noise. In contrast, our study found no such association between the IPL and AzBio sentence scores under any listening condition, regardless of how the IPL was quantified. We identified three key methodological differences in IPL quantification between Schvartz-Leyzac et al. [21] and our study. First, the number of sweeps included in the averaged eCAP responses differed. Schvartz-Leyzac et al. [21] used 50–100 sweeps, whereas we used 400 sweeps, potentially improving the SNR in our data. Second, there were differences in the designated latency windows for identifying the N1 and P2 peaks. Schvartz-Leyzac et al. [21] reported using latency windows of 150–300 μs for N1 and 400–600 μs for P2. However, we noted a discrepancy between these reported time windows and the IPL values shown in their Figures 5 and 6. Based on their defined latency windows, all IPLs should fall within 450 μs or less, yet several IPLs reported in their study exceeded 450 μs, with one even surpassing 500 μs. The reason for this inconsistency is unclear. In our study, broader latency windows were used (i.e., 150–600 μs for N1 and 400–1000 μs for P2) to better accommodate variabilities in eCAP morphology. Finally, Schvartz-Leyzac et al. [21] did not use any rounding up process to control for the multi-peak issue.

To determine whether the discrepancies between our findings and those of Schvartz-Leyzac et al. [21] were attributable to methodological differences, we reanalyzed our data using the time windows reported in their Methods section to identify N1 and P2 peak latencies. Specifically, IPLs were recalculated for averaged eCAPs across both the first 50 sweeps and the full set of 400 sweeps, without applying any rounding-up procedure. We then reexamined the associations between IPL and psychophysical GDTs, participant age, and AzBio sentence scores obtained under various listening conditions in adult CI users. Supplemental Digital Content 1 shows the association between the IPL, psychophysical GDT and AzBio sentence scores measured in quiet and in noise. For the IPL calculated for the averaged eCAP across 50 sweeps, five participants showed equal IPLs at the two electrode locations with different psychophysical GDTs. Smaller GDTs measured at the electrodes associated with shorter IPLs in five participants. An opposite pattern was observed in four participants. When using IPLs derived from 400-sweep averages, nine participants had equal IPLs at two electrode locations with different GDTs, effectively ruling out the IPL as a reliable indicator of GDT. In addition, no associations were found between the IPL and AzBio sentence scores under any listening condition, regardless of the number of sweeps used in eCAP averaging. The MLR results supporting this observation are presented in Table 2. Supplemental Digital Content 2 shows IPLs measured at different electrode locations as a function of participant age. These data revealed no correlation between IPL and age at any electrode location, regardless of the sweep count used in the averaged eCAP. This lack of association is further supported by Pearson correlation results, as indicated in each panel. In summary, our reanalysis showed that the discrepancies between our findings and those of Schvartz-Leyzac et al. (2025) cannot be explained by the three methodological differences identified earlier. At present, the factors accounting for this inconsistency remain unknown.

### The inter-trial variability of the interpeak latency of the eCAP

The robust correlation between the VIL and the PLV indicates that both measures likely assess common underlying biological mechanisms that produce temporal jitter in the single-sweep eCAP waveform, as was simulated in Figure 8. This interpretation is further supported by the similarity in their results, including the effects of electrode location and their associations with temporal resolution and speech perception outcomes, as reported in our previous studies [17,18]. Consequently, these findings are not repeated or discussed in detail in the current report. Instead, we summarize the advantages and limitations of each measure below.

#### The VIL

##### Pros

This measure can be applied across CI devices regardless of the manufacturer. The computational method is straightforward and easy to understand. No parameter adjustment is needed when testing patients with CIs from different manufacturers. The VIL is only sensitive to the variability in peak latencies across trials, in contrast to the IPL’s additional dependence on the mean peak latencies (i.e., latency differences that are common across trials).

##### Cons

As with IPL, VIL can be affected by the presence of multiple peaks in eCAP waveforms, which can compromise precision. In the results presented in this paper, the prevalence of multiple peaks is greater in the VIL method than the IPL approach, because the VIL method is based on single sweeps and therefore does not average out the recording noise. Additionally, our analysis of the eCAP recording-noise spectrum indicates that the recording noise has a greater impact on the VIL than on the PLV. The varying recording noise levels across CI devices may hinder the consistent application of VIL as a cross-platform metric.

#### The PLV

##### Pros

The PLV is not affected by the ambiguity introduced by multiple peaks in eCAP waveforms and has a better effective SNR than the IPL and VIL. The PLV is a well-established concept for assessing neural synchrony in literature and is only sensitive to variability in the eCAP waveform across trials, not the basic shape of the single-sweep waveform arising from factors that are common across trials, in contrast to the IPL.

##### Cons

Parameters used in mathematical computations need to be adjusted based on the sampling rate of CI devices from each manufacturer. Current parameter settings are optimized for Cochlear™ Nucleus® devices only. Specific parameters for other CI systems remain undetermined. This limitation complicates direct comparison of PLV results between users of different CI devices.

## CONCLUSIONS

There is no compelling evidence from our data or simulations supporting the IPL as a reliable indicator of neural synchrony in the CN. However, its inter-trial variability could be used as an alternative to the PLV to evaluate neural synchrony of the CN in CI users. The precision of this measure is affected by the sampling rate and recording noise of CI devices.

## Data Availability

All data used in the present study are available upon reasonable request to the authors.

