## Supplemental Figure 1 for "Evaluating the N1-P2 interpeak latency of the eCAP and its inter-trial variability as potential indicators of neural synchrony in the cochlear nerve of cochlear implant users"

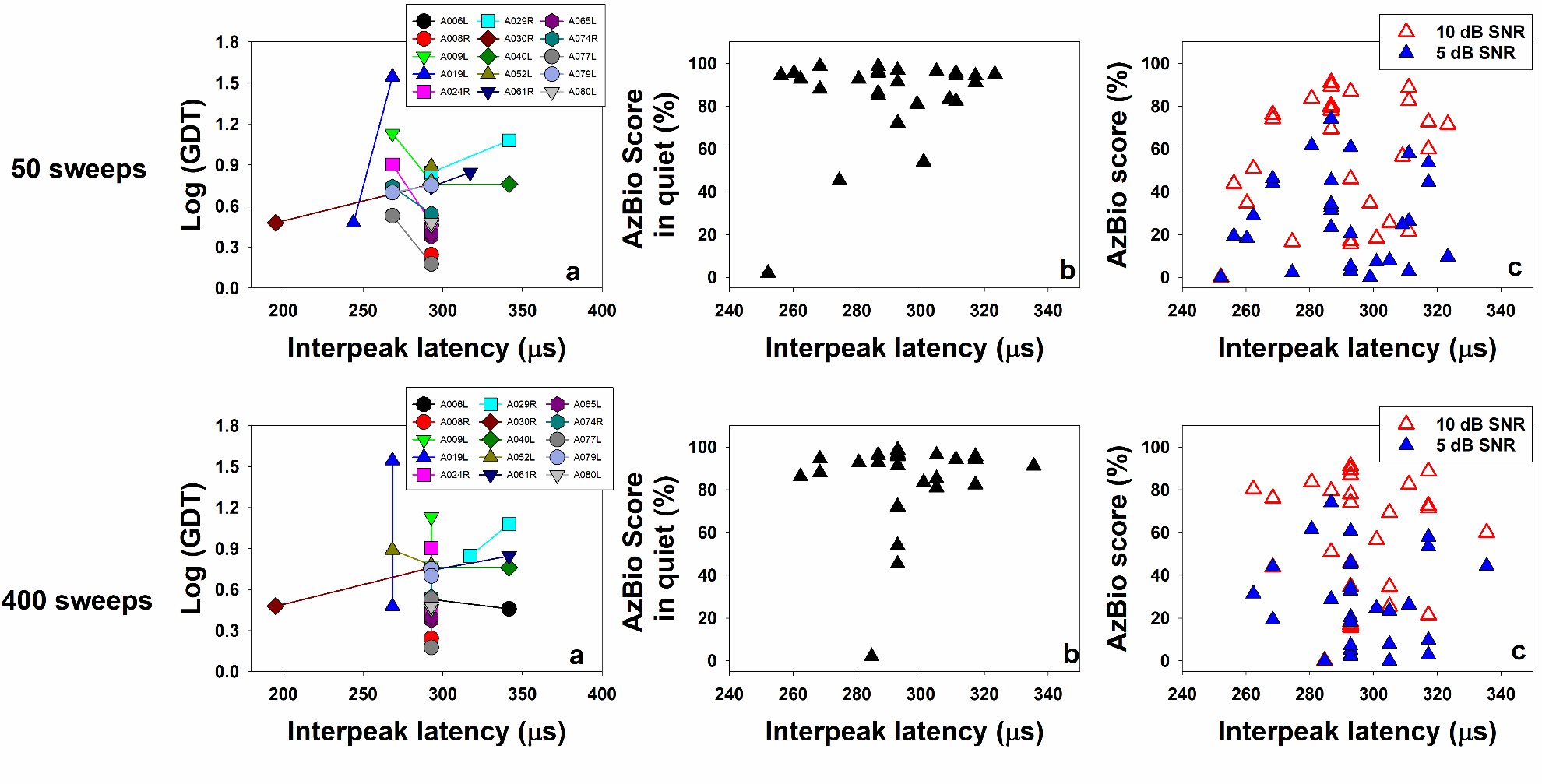

**Figure A**. The associations between interpeak latencies quantified using the designated latency window reported in Schvartz-Leyzac et al. (2025) and auditory perception outcomes. Panels listed in the top and the bottom rows represent the results averaged across 50 and 400 sweeps, respectively. In each row, panel (a) displays interpeak latencies (IPLs) and psychophysical gap detection thresholds (GDTs) measured at two electrode locations in each of 15 adult participants. Lines connect the data measured at the two electrode locations tested in the same ear. Panel (b) in each row shows scatter plots of the association between the IPL and AzBio sentence scores measured in quiet in adult participants tested in this study. Panel (c) in each row shows scatter plots of the association between the IPL and AzBio sentence scores measured in the two noise conditions in the same group of adult participants. Each symbol indicates the result measured in one participant in panels (b) and (c).
