## Supplemental Figure 2 for "Evaluating the N1-P2 interpeak latency of the eCAP and its inter-trial variability as potential indicators of neural synchrony in the cochlear nerve of cochlear implant users"

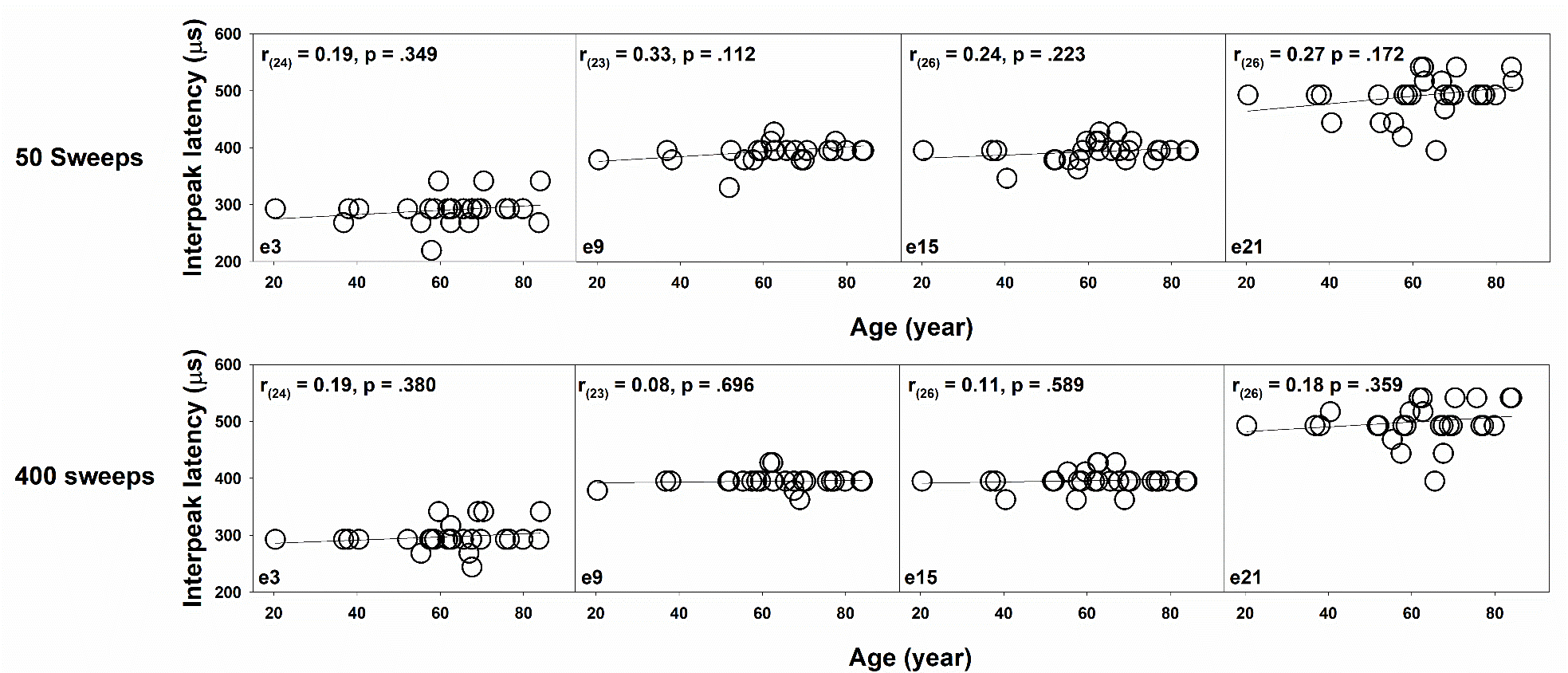

**Figure B**. The associations between interpeak latencies quantified using the designated latency windows reported in Schvartz-Leyzac et al. (2025) and participant age. Panels listed in the top and the bottom rows represent the results averaged across 50 and 400 sweeps, respectively. Each panel represents the results measured at one electrode location. Each symbol indicates the result from one participant. Solid lines represent the results of linear regression. The results of Pearson correlation tests are indicated in each panel.
